# Influenza infection, Acute myocardial Infarction, Flu Shot during COVID-19 Pandemic in US population. A Review of Literature

**DOI:** 10.1101/2021.07.14.21260549

**Authors:** Nischit Baral, Nirajan Nayak

## Abstract

Influenza is a major cause of hospitalization in all age groups but can cause more severe infections in specific high-risk population. Novel Corona Virus Disease 2019 (COVID-19) pandemic and Influenza virus infection cause similar illness and coexist. Cardiovascular complications due to influenza are important causes of morbidity and mortality in the US, especially in the elderly population (aged more than 65 years). Acute Myocardial Infarction (AMI) is the most serious among the cardiovascular causes of mortality following the attack of influenza, mainly in patients with various co-morbidities like pre-existing coronary artery disease (CAD), diabetes mellitus (DM), hypertension (HTN), and heart failure (HF). We have reviewed the association between influenza virus infection and AMI and extrapolated the beneficial effects of influenza vaccine in preventing AMI and its grave consequences. We have also highlighted about the importance of flu shot during the COVID-19 pandemic.

## Introduction

Influenza complications are important causes of morbidity and mortality in many parts of the world, especially in the elderly population (aged>65 years). This seems to be on an increasing trend with the increase in life expectancy in the aging population^1.^ Acute Myocardial Infarction (AMI) is the leading cardiovascular complication of influenza virus infection more importantly in the elderly and the high-risk individuals^2-4^. Novel Corona Virus Disease 2019 (COVID-19) is an on-going pandemic and Influenza virus infection may coexist with COVID-19. According to the data from Center of Disease Control, US from October 1, 2019, through April 4, 2020, there were 39,000,000- 56,000,000 flu illnesses, 410,000- 740,000 laboratory-confirmed-flu hospitalizations, and 24,000- 62,000 flu deaths. The majority (58%) of influenza-associated hospitalizations were in the elderly age group^5^. Younger patients (age < 65 years)were at risk of influenza-related complications only in the setting of high-risk factors such as immune-compromised states, coronary artery disease (CAD), heart failure (HF) exacerbations, hypertension (HTN) and diabetes mellitus (DM)^6^. COVID-19 pandemic is still a global problem with new Delta variant infecting large population in the US as well all over the world. The role of COVID-19 vaccine as well as flu vaccine is more important during these times as per the recommendations from CDC^5^.

### Objective

The purpose of this article is to provide an updated review on influenza/flu illness and AMI and highlight the role of flu vaccination in times of COVID-19 pandemic.

## Materials and methods

### Data extraction

We searched Medline and Embase using relevant Medical Subject Headings (MeSH) termed influenza or influenza virus or flu or Corona virus, COVID-19, Novel Corona Virus, SARA-CoV-2, flu vaccine, and myocardial infarction or acute myocardial infarction or STEMI/ACS or heart attack literature published within the last five years with additional filters of human studies and customized articles. The titles and abstracts of all results were reviewed and studies were selected for full-text analysis according to their eligibility criteria. Figure 1.

**Figure 1:**
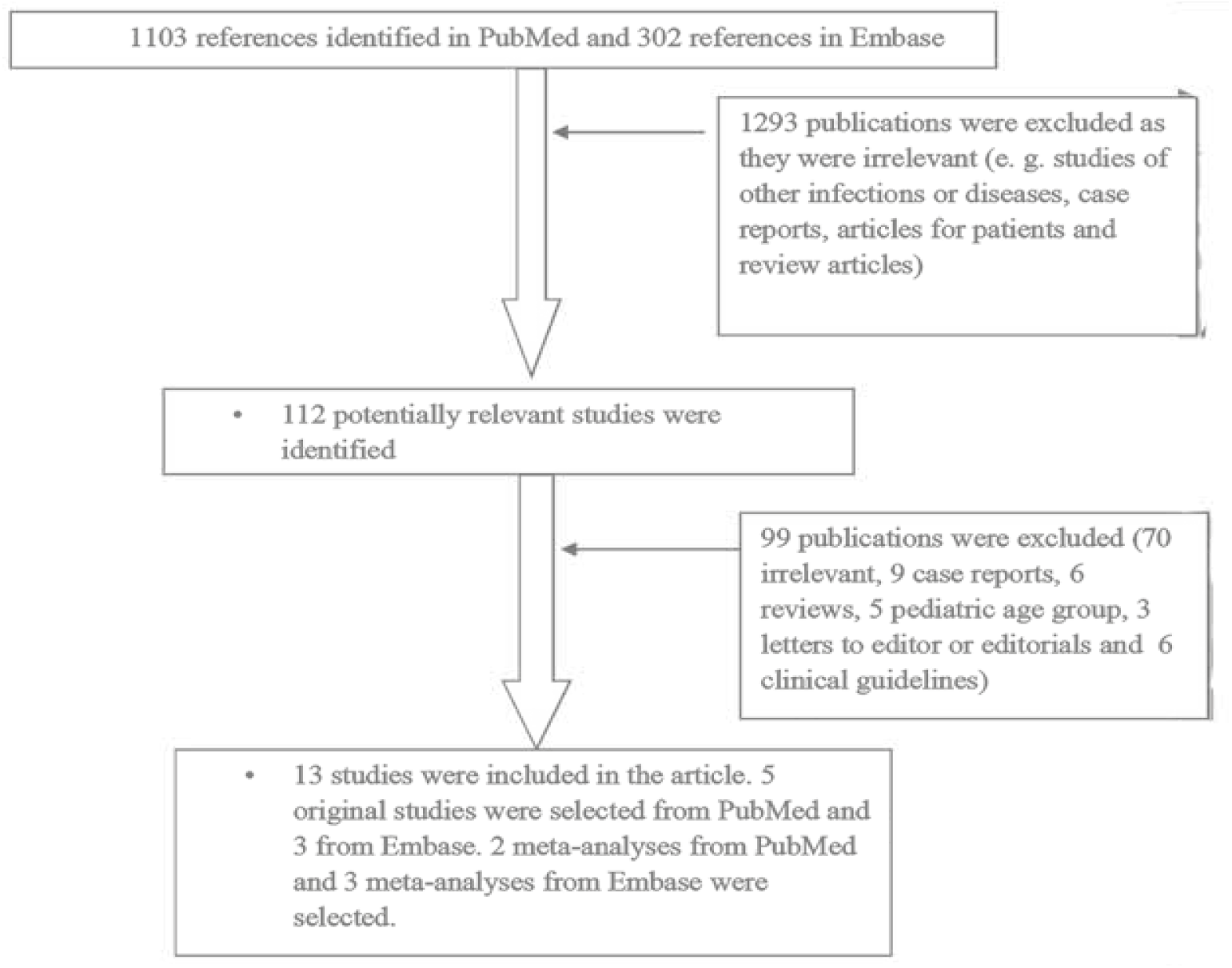
Flow Diagram of Included Studies

#### Eligibility criteria

We included human studies on AMI, COVID-19 and influenza or influenza vaccination in adults (age more than or equal to18 years) for full-text analysis. We excluded review articles and case reports.

#### Quality assessment and data extraction

Authors NB and NN independently performed the study selection, data extraction and quality assessment.in the prevention of AMI associated hospitalization and death.

## Results and Discussion

### Prevalence of influenza triggered AMI

Influenza is a trigger for AMI and ischemic heart disease (IHD) particularly in the elderly population^4, 7-9.^ Warren-Gash et al^10^. performed a multicentre prospective time series study in England, Wales and Hong Kong and found that influenza was associated with increased AMI related hospitalization and death, suggesting this infection as an independent risk factor for AMI^10^. This is further supported by an observational study by Song et al. who too reported that elderly patients suffering from influenza and having other comorbidities such as DM, HF, pre- existing CAD and HTN had higher in-hospital cardiac mortality (fatal cardiac arrest and ventricular arrhythmias) as compared to patients without these risk factors^11^.

A similar association between infection due to influenza virus and AMI was documented by Pearce et al.8 and Kwong et al.^4^ substantiated that the influenza virus was a stronger risk factor for the development of AMI than any other virus causing acute respiratory tract infection. A time series study by Nguyen et al. found that there was a significant increase in all cause cardiovascular mortality and AMI mortality during the influenza outbreak^1^

Guan and colleagues in the recent past stated that cases of AMI were more likely to have positive IgG antibodies to influenza virus A and B as compared to controls; an observation which supported the hypothesis that previous influenza virus infection had a role to play in the development of atherosclerosis, eventually triggering the occurrence of AMI^13^.

### Pathogenesis of post-influenza AMI

The precise mechanism for the development of AMI following influenza virus infection is unclear. However, Naghavi et al. proposed that the virus promoted atherosclerotic plaque formation by inducing inflammation, smooth muscle cell proliferation and fibrin deposition. Histological analysis from the aortic sections of apolipoprotein E-deficient mice injected with influenza A virus showed marked proliferation of intimal epithelial cells along with smooth muscle cells of tunica media, which were filled with macrophages and T lymphocytes, quite suggestive of atherosclerosis^14^.

Notwithstanding the above, the exact link between the formation of atherosclerotic plaque and influenza infection has not been clearly delineated. MacIntyre et al. recently proposed that influenza potentiated the release of inflammatory cytokines which could give rise to a prothrombotic state^15^.Further, influenza might have a direct effect on the heart, producing local inflammatory changes, as evidenced by histopathological and molecular studies on influenza-infected mice^16^. Thus, a series of studies conducted in the recent past, at various centers supported the above-mentioned hypothesis and these data have been presented vide Table 1.

**Table 1:**
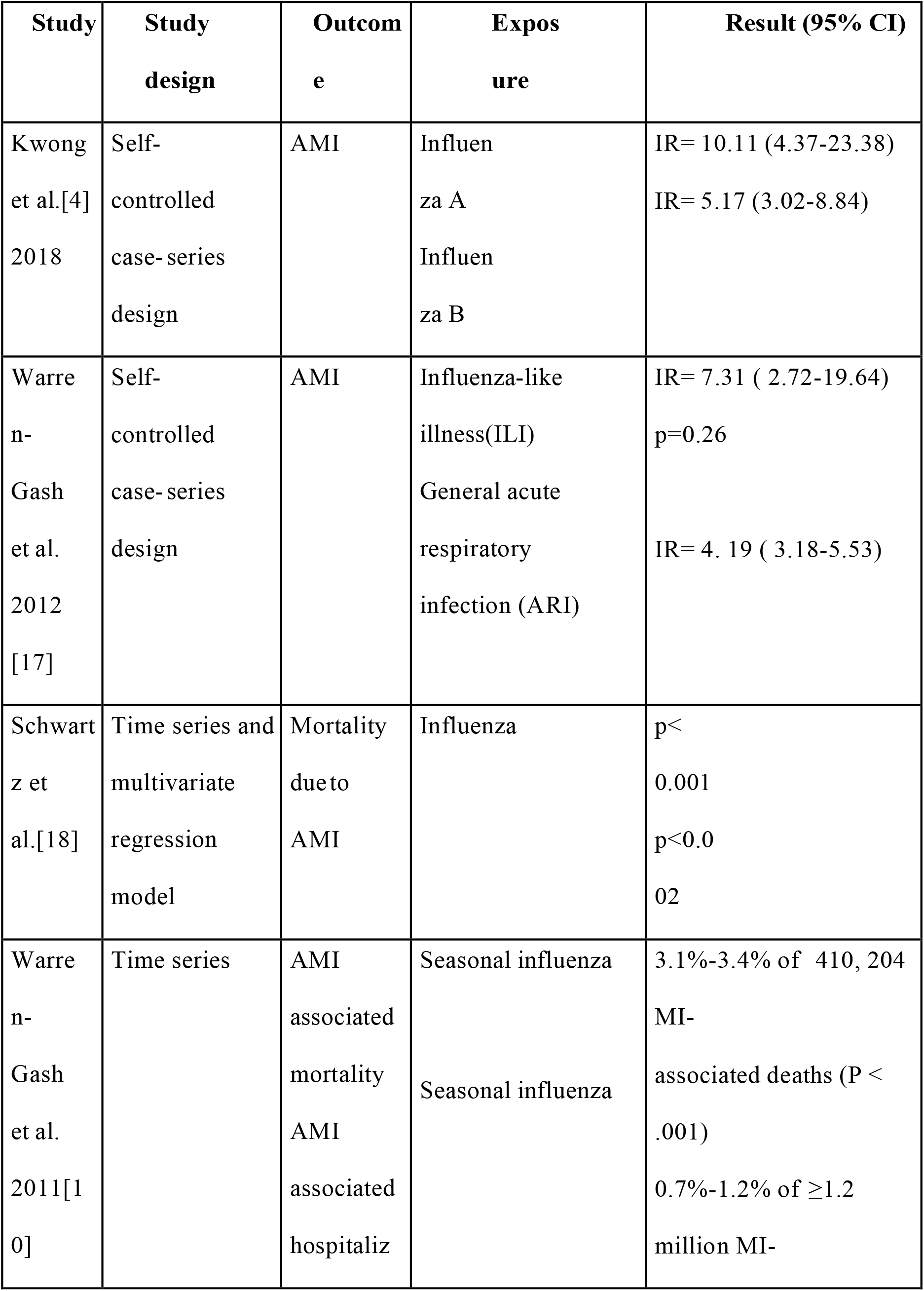
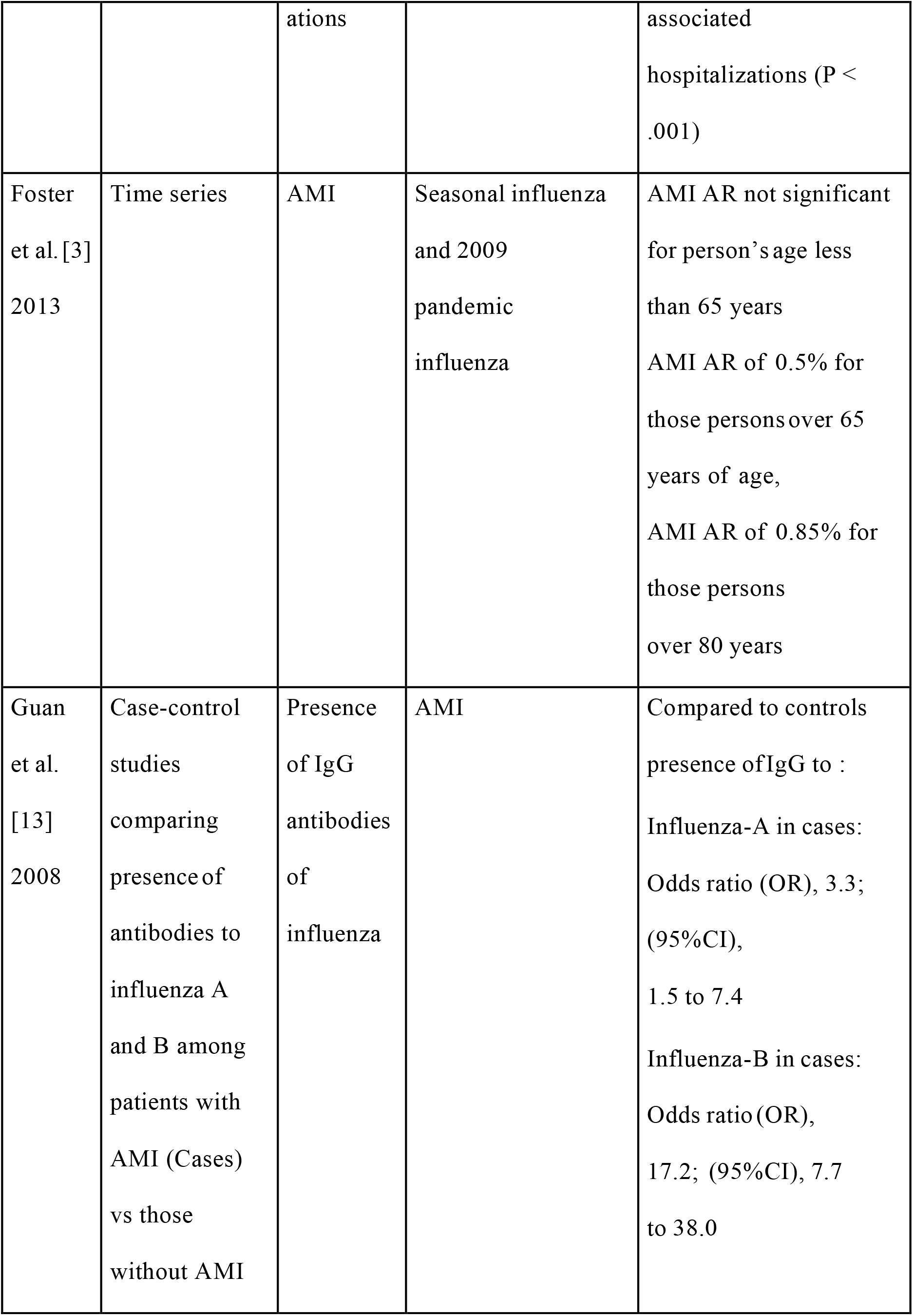
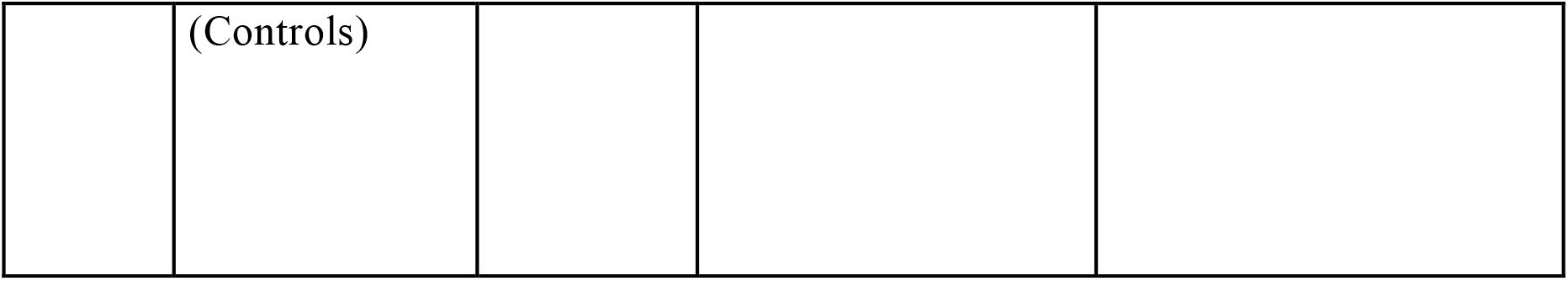
Summary of reports depicting the role of influenza as a trigger for AMI. IR: Incidence Ratio CAD: Coronary Artery Disease AR: Attributable Risk

### COVID -19 and Cardiovascular Risk

As per the CDC reports, COVID-19 is associated with various cardiovascular diseases similar to Flu. The Corona virus has many unknown etiologies that can cause cardiovascular disease. On the other hand, flu virus is associated with similar disease^5^. However as per CDC, flu virus causes mild illness in comparison to COVID-19. But Flu virus is also associated with Cardiovascular infection^5, 19, 20^. A study by Nayak et al highlights that, to prevent AMI hospitalizations and mortality following an attack of influenza, prompt administration of influenza vaccine is needed^20^. Huang et al. conducted a cohort study and showed a significant reduction in the risk of ischemic heart disease in elderly patients receiving influenza vaccine^19,20^. A systematic review by Claret al. also showed that AMI was significantly reduced after influenza vaccination^20^. In a prospective randomized open-ended study by Phrommintikul et al., influenza vaccination was shown to decrease major cardiovascular hospitalizations due to AMI. However, no significant difference was found in the incidence of cardiovascular deaths among vaccine recipient and non-recipient groups21.This is further supported in a meta-analysis by Udell et al, however, the difference is statistically non-significant and further studies are warranted to prove the benefit of influenza vaccine in the prevention of cardiovascular mortality, especially in high-risk patients with previous acute coronary syndrome22. However, vaccine, by preventing influenza and its associated hospitalizations, in a way could prevent the possibility of AMI triggered by the mechanisms discussed above and also might prevent the associated AMI related mortality.

Just as influenza vaccine prevents the AMI and its associated hospitalizations, COVID-19 vaccine may prevent AMI and associated Cardiovascular hospitalizations20, 23. We need more studies to comment on these topics. Vaccination should be encouraged to prevent cardiovascular complications and mortality especially among high-risk and elderly patients, in these challaging times of COVID-19 ^5, 20-28^. Vaccination also prevents from influenza myocarditis^28^.

## FLU SHOT IN TIMES OF COVID-19

According to CDC, flu shot/influenza vaccination has been deemed to be more important in times of COVID-19 due to various reasons^5^. Flu shots protect us from getting flu-like illness which has similar presentation as COVID-19. There may be difficulty in diagnosis COVID-19 due to flu-like-illness. There are no any studies that show increased risk of getting COVID-19 from Flu shots^5^. A recent systematic review showed the role of aspirin in improving mortality from COVID-19^27^. Though, we lack any evidence on the role of flu-shot in prevention of COVID-19, flu shot in times of COVID-19 is recommended, as co-infection with COVID-19 and Influenza may occur^5^.

## CONCLUSION

Both Flu-shot and COVID-19 vaccination are important during this COVID-19 pandemic.

## Data Availability

Please email at

